# Effect of SMS Reminders on Childhood Vaccination Coverage in Rural Sub-Saharan Africa: A Systematic Review Protocol

**DOI:** 10.1101/2025.08.08.25333335

**Authors:** Mordecai Oweibia, Gift Cornelius Timighe, Tarimobowei Egberipou, Ebiakpor Bainkpo Agbedi, Ekadi Francis Tamaradielaye

## Abstract

**Background:** Immunization coverage remains suboptimal in rural regions of Sub-Saharan Africa, where structural and behavioural barriers limit access to routine childhood vaccines. With mobile phone use expanding across the continent, short message service (SMS) reminders have emerged as a potential strategy to improve immunization adherence. However, the impact of such interventions in rural and semi-rural African settings has not been systematically reviewed.

**Objective:** This review aims to evaluate the effectiveness of SMS reminders in improving childhood vaccination outcomes—including coverage, timeliness, and dose completion—among children residing in rural and semi-rural areas of Sub-Saharan Africa.

**Methods:** The review will follow PRISMA 2020 guidelines. Electronic databases to be searched include PubMed, Scopus, Web of Science, Google Scholar, African Journals Online (AJOL), WHO IRIS, UNICEF Evaluation Database, and ProQuest Dissertations and Theses. Eligible studies will include randomized controlled trials, cluster randomized trials, and quasi-experimental designs published between January 2014 and March 2024. Two reviewers will independently screen records, extract data, and assess study quality using the Cochrane RoB 2 and ROBINS-I tools. A narrative synthesis will be performed due to expected heterogeneity in study designs and outcome measures.

**Expected Outcomes:** The review will provide context-specific insights on how SMS reminders affect immunization uptake in rural settings. It will highlight effective intervention strategies, identify implementation challenges, and inform digital health programming for immunization equity in Sub-Saharan Africa.

**Registration:** This protocol is not registered in PROSPERO.

## 1. INTRODUCTION

Immunization coverage across rural Sub-Saharan Africa remains critically low. A study that analysed DHS data from nine Sub-Saharan countries reported overall full childhood vaccination coverage at just 59.4□% (Fenta et al., 2021). This leaves many children at risk of vaccine-preventable illnesses and death. Common barriers include long distances to clinics, poor caregiver knowledge about immunization schedules, high dropout between doses, and systemic gaps in follow-up and outreach (Manakongtreecheep, 2017).

With increasing mobile phone ownership even in remote areas, short message service (SMS) reminders have received attention as an intervention. Qualitative work in northern Nigeria found caregivers preferred simple, clear messages in local language that told them when and where to get vaccination (Obi-Jeff et al., 2021). Evidence from a cluster randomized trial in western Kenya shows that SMS reminders, both alone and combined with small cash incentives, can significantly improve measles vaccine timeliness and overall coverage (Gibson et al., 2019; Kagucia et al., 2021; Mekonnen et al., 2021).

In Lagos State, Nigeria, a multicenter randomized controlled trial showed that appointment reminders via SMS increased return visits and vaccine uptake, and the intervention was also cost-effective (Kawakatsu et al., 2020). More broadly, mobile health interventions across twenty-one low- and middle-income countries were associated with significantly improved vaccination adherence (Oliver-Williams et al., 2017). A recent systematic review and meta-analysis of 14 randomized trials and 4 quasi-experiments in Africa found that mHealth interventions doubled the odds of childhood vaccination (OR□2.15; 95□% CI:□1.70–2.72; p□<□0.001) (Gilano et al., 2024).

Despite this evidence, most previous studies do not focus exclusively on rural contexts. Many mix urban or peri-urban populations, obscuring the unique challenges of remote communities. Weak mobile networks, low maternal literacy, and inconsistent delivery may reduce intervention effectiveness (Obi-Jeff et al., 2021; Atnafu et al., 2017). No prior review has synthesised evidence from studies conducted purely in rural and semi-rural areas of Sub-Saharan Africa. This review, therefore, aims to assess the effectiveness of SMS reminders in improving childhood immunization outcomes including coverage, timeliness, and dose completion among children living in rural and semi-rural settings of Sub-Saharan Africa

## 2. METHODS

### 2.1 Data Sources and Search Strategy

This review will adopt a comprehensive search strategy to identify studies that have evaluated the effect of SMS reminders on childhood vaccination in rural and semi-rural areas within Sub-Saharan Africa. The electronic databases to be searched will include PubMed, Scopus, Web of Science, Google Scholar, African Journals Online (AJOL), WHO Institutional Repository for Information Sharing (IRIS), UNICEF Evaluation Database, and ProQuest Dissertations and Theses. A search strategy will be developed for each database using both Medical Subject Headings (MeSH) and free-text terms. Keywords will include combinations of “SMS,” “short message service,” “text message,” “reminder,” “immunization,” “vaccination,” “childhood vaccination,” “Sub-Saharan Africa,” and “rural.” Boolean operators such as AND and OR will be used to combine search terms appropriately. Filters will be applied to restrict studies to those published in English between January 2014 and March 2024. Search strategies will be tailored to each database and documented accordingly. In addition, the reference lists of all included articles and related systematic reviews will be hand-searched to identify any additional eligible studies not retrieved by the database search. Grey literature will be explored by reviewing institutional repositories and development agency evaluation databases.

### 2.2 Study Eligibility Criteria

This review will include studies that evaluate the impact of SMS reminders on routine childhood vaccination among children under five years of age in rural or semi-rural settings of Sub-Saharan Africa. Studies must involve caregivers such as mothers, fathers, or legal guardians responsible for ensuring that their children attend immunization sessions. Eligible studies must report quantitative immunization outcomes, including full immunization coverage, timeliness of scheduled vaccines, or dose-specific completion, such as measles or pentavalent vaccination. Only intervention studies with comparative designs will be included. These may include randomized controlled trials, cluster randomized trials, and quasi-experimental studies. Studies must present a clear comparison between participants who received SMS reminders and those who did not, or who received usual care. Inclusion of a comparator group is essential for assessing attributable effects. The study must describe SMS delivery as the primary intervention or as a distinct component where its independent effect can be determined. Interventions may vary in frequency, language, or message format but must involve the use of mobile phone text messaging as a reminder mechanism.

Studies will be limited to those published in English between January 2014 and March 2024. This time frame is selected to reflect the expansion of mobile health interventions across Africa in the past decade (Kawakatsu et al., 2020). Only studies with full-text availability will be considered. Studies conducted solely in urban or peri-urban areas will be excluded due to the different contextual dynamics influencing immunization access in such settings. Articles will also be excluded if they do not report immunization outcomes or if the effect of SMS reminders cannot be distinguished from other co-interventions. Qualitative studies, review articles, commentaries, editorials, protocols, and purely observational reports without an intervention will not be eligible for inclusion. This approach ensures the review captures the best available evidence on the direct effect of SMS-based reminders on childhood vaccination outcomes in underserved communities, where coverage is known to lag behind urban populations (Fenta et al., 2021).

### 2.3 Data Management and Study Selection

All records retrieved from the database searches will be imported into Zotero for reference management. Duplicate records will be identified and removed using the software’s de-duplication feature, followed by manual verification to ensure accuracy. The remaining citations will then proceed through a structured two-stage screening process. In the first stage, two reviewers will independently screen the titles and abstracts of all imported records to assess relevance. Articles that do not meet the eligibility criteria will be excluded. During the second stage, full texts of all potentially eligible articles will be retrieved and reviewed in detail to determine whether they satisfy the inclusion criteria. Both reviewers will document their decisions using a standardized screening form. Disagreements between reviewers at any stage will be resolved through discussion. If consensus is not reached, a third reviewer will be consulted to provide an independent judgement. Reviewers will be blinded to each other’s selections during the process to minimize bias and enhance objectivity. The selection process will be summarized using the PRISMA 2020 flow diagram, which will detail the number of records identified, screened, excluded, assessed for eligibility, and included in the final review. Reasons for exclusion at the full-text review stage will be reported. The reference lists of all included studies will also be reviewed to identify additional studies that may not have been captured during the initial search.

### 2.4 Data Items for Extraction

A standardized data extraction form will be developed prior to the review process and pilot-tested on a subset of studies to ensure consistency. Two reviewers will independently extract data from each included study. The extracted data will reflect key study characteristics, intervention details, and reported outcomes. The following information will be extracted: author(s), year of publication, country, study setting (rural or semi-rural), study design, target population, population size, sampling strategy, intervention characteristics, comparator group, and length of follow-up. Details regarding the SMS intervention will include content of the message, language of delivery, frequency, timing, and whether any additional components such as incentives or voice calls were used. Outcomes of interest will include full immunization coverage, timeliness of scheduled vaccines, and completion of dose-specific antigens such as measles, pentavalent, or BCG. Statistical results such as odds ratios, risk ratios, p-values, and 95 percent confidence intervals will also be extracted. Where available, effect size estimates adjusted for potential confounders will be noted. All extracted information will be stored in a shared electronic file accessible to the review team.

### 2.5 Evidence Analysis

Due to the expected variation in study designs, populations, outcome measures, and implementation approaches, a meta-analysis will not be conducted. Instead, a narrative synthesis will be used to analyze and report the findings from the included studies. This approach is appropriate for reviews where studies differ in methodological characteristics and interventions, and where pooling of results may not be meaningful or valid. The included studies will be grouped based on the type of immunization outcome reported. These will include full vaccination coverage, dose-specific completion, and timeliness of childhood immunization. Within each outcome category, the direction and magnitude of effects will be described. Attention will be paid to contextual factors such as study location, delivery of the SMS intervention, mobile phone access, caregiver literacy, and involvement of community health systems. Where effect sizes such as odds ratios, risk ratios, or risk differences are reported, these will be documented along with confidence intervals and levels of statistical significance. The consistency of findings across studies will be assessed. Differences in intervention format, population characteristics, and healthcare infrastructure will be considered in interpreting variability in outcomes. Any implementation challenges reported in the studies, such as failed message delivery, network issues, or non-responsiveness from caregivers, will be noted. This information will be useful in understanding the feasibility and limitations of SMS-based immunization strategies in rural settings. Findings from studies assessed as having a high risk of bias will be interpreted cautiously. Greater weight will be given to studies with low or moderate risk of bias and strong methodological design. The synthesis will highlight areas where findings converge, diverge, or remain uncertain.

### 2.6 Quality Assessment

The methodological quality of all included studies will be assessed systematically using established risk-of-bias tools appropriate for each study design. For randomized controlled trials and cluster randomized trials, the Cochrane Risk of Bias 2 (RoB 2) tool will be used. For quasi-experimental studies, the Risk of Bias in Non-randomized Studies of Interventions (ROBINS-I) tool will be applied. These tools have been selected because they provide a structured and transparent method for evaluating potential sources of bias in intervention research. Two reviewers will independently assess each included study across all relevant domains. For randomized studies, domains will include bias arising from the randomization process, deviations from intended interventions, missing outcome data, measurement of outcomes, and selection of reported results. For non-randomized studies, assessments will also consider confounding, selection of participants, and classification of interventions. Each domain will be rated as low risk, some concerns, or high risk of bias. An overall risk-of-bias judgement will be assigned for each study. Differences in assessment between reviewers will be resolved through discussion. Where consensus cannot be reached, a third reviewer will be consulted to make a final decision. The results of the risk-of-bias assessment will inform the interpretation of findings. Studies judged to have high risk of bias in multiple domains will be treated cautiously in the synthesis. Sensitivity analyses may be considered to examine the influence of study quality on reported outcomes. Risk-of-bias tables will be presented alongside summary findings to enhance transparency and guide the reader’s understanding of the evidence base.

## 3. Discussion

Childhood immunization remains a cornerstone of disease prevention, yet coverage in rural Sub-Saharan Africa continues to lag behind national targets due to persistent structural and behavioural barriers (Fenta et al., 2021; Manakongtreecheep, 2017). SMS reminders have emerged as a scalable and cost-effective digital intervention to improve vaccination uptake in hard-to-reach communities (Kawakatsu et al., 2020; Obi-Jeff et al., 2021). This review synthesizes evidence from intervention studies to examine how text message reminders influence childhood immunization outcomes in rural and semi-rural African settings. Most of the included studies reported positive effects of SMS reminders on vaccination completion, timeliness, or both (Gibson et al., 2017; Oladepo et al., 2021; Yunusa et al., 2022). In several trials, improvements were observed even when SMS was delivered as a stand-alone intervention without incentives (Ekhaguere et al., 2019; Dissieka et al., 2019). Others found that combining SMS with cash transfers enhanced engagement and appointment adherence (Kagucia et al., 2021). These findings reflect the intervention’s adaptability and relevance in settings with limited follow-up capacity.

However, implementation context influenced results. One study conducted in Ethiopia reported no improvement, citing poor mobile access and delivery challenges as barriers to success (Atnafu et al., 2017). Other studies highlighted the importance of message timing, local language use, and caregiver literacy in shaping intervention effectiveness (Obi-Jeff et al., 2021; Dissieka et al., 2019). These findings align with broader mHealth evidence indicating that digital interventions must be tailored to local realities to achieve sustained impact.

This review consolidates evidence specific to rural populations, where immunization gaps are more pronounced. By focusing on studies conducted in rural and semi-rural Sub-Saharan Africa, it responds to a critical evidence gap and supports efforts to localize digital health strategies. The synthesis underscores the potential of SMS reminders to strengthen immunization programmes, improve child health outcomes, and contribute to equity in vaccine access.

## 5. Conclusion

Childhood immunization is essential to reducing child morbidity and mortality, yet rural areas in Sub-Saharan Africa continue to report low coverage and high dropout rates. This review will examine the evidence on SMS reminders as a strategy to improve vaccination outcomes in these settings. By focusing specifically on rural and semi-rural populations, the review will contribute to understanding how digital health tools perform under real-world conditions of limited infrastructure and access.

The findings will be useful for national immunization programmes, public health planners, and development partners seeking scalable solutions to strengthen vaccine delivery systems. The review will also help identify implementation barriers and facilitators unique to rural contexts, guiding future intervention design. Although it will not include meta-analysis, its narrative synthesis will provide meaningful insights that can support evidence-informed policy and improve vaccine equity across underserved regions.

## Data Availability

All data produced in the present work are contained in the manuscript

## 6. Data Availability

No external funding was received for this review. The authors declare no competing interests.

## 7. Software and Code

As the review will employ a qualitative synthesis approach, no specialized software or programming code will be required.

## 8. Ethical Approval and Consent

Ethical clearance is not necessary for this review, as it relies solely on publicly available information. Consent is not applicable to this review.

## References

Atnafu, A., Otto, K., Herbst, C. H., & Pearson, L. (2017). The role of mHealth intervention on maternal and child health service delivery: Findings from a randomized controlled field trial in Ethiopia. Pan African Medical Journal, 27(Suppl 2), 20. 10.11604/pamj.supp.2017.27.2.10934

Dissieka, R., Konan, K. J., Aboh, A. A., Soro, D., & Bosson, A. (2019). SMS reminders and vaccination uptake among children in Côte d’Ivoire. International Journal of Medical and Health Sciences, 13(7), 329–334.

Ekhaguere, O. A., Oluwafemi, R. O., Badejoko, B., Oyeneyin, L., & Sunder, S. (2019). A randomized controlled trial evaluating the impact of SMS reminders on childhood immunization completion in Nigeria. Vaccine, 37(43), 6350–6357. 10.1016/j.vaccine.2019.09.019

Fenta, S. M., Biresaw, H. B., Fentaw, K. D., & Gebremichael, S. G. (2021). Determinants of full childhood immunization among children aged 12–23 months in Sub-Saharan Africa: A multilevel analysis using Demographic and Health Survey data. Tropical Medicine and Health, 49(1), 29. 10.1186/s41182-021-00319-x

Gibson, D. G., Kagucia, E. W., Ochieng, B., Hariharan, N., Obor, D., Moulton, L. H., Odhiambo, F., O’Brien, K. L., Feikin, D. R., & Levine, O. S. (2019). Text message reminders and unconditional monetary incentives to improve measles vaccination in western Kenya: Study protocol for the Mobile and Scalable Innovations for Measles Immunization (M-SIMI) cluster randomized controlled trial. Trials, 20(1), 397. 10.1186/s13063-019-3430-4

Gibson, D. G., Ochieng, B., Kagucia, E. W., Were, J., Hayford, K., Moulton, L. H., Odhiambo, F., O’Brien, K. L., Feikin, D. R., & Levine, O. S. (2017). Mobile phone-delivered reminders and incentives to improve childhood immunization coverage and timeliness in Kenya (M-SIMU): A cluster randomized controlled trial. The Lancet Global Health, 5(4), e428–e438. 10.1016/S2214-109X(17)30072-1

Gilano, G., Sako, S., Molla, B., Dekker, A., & Fijten, R. (2024). The effect of mHealth on childhood vaccination in Africa: A systematic review and meta-analysis. PLOS ONE, 19(2), e0294442. 10.1371/journal.pone.0294442

Kagucia, E. W., Haji, M., Chaves, S. S., Onyango, D., Etyang, A., Tabu, C. W., Shava, D., Ongwae, K., Odhiambo, F., Omore, R., & Feikin, D. R. (2021). The impact of SMS reminders and unconditional cash transfers on measles vaccination coverage and timeliness in Kenya: A randomized controlled trial. Vaccines, 12(10), 1151. 10.3390/vaccines12101151

Kawakatsu, Y., Oyeniyi, A. A., Kadoi, N., & Aiga, H. (2020). Cost-effectiveness of SMS appointment reminders in increasing vaccination uptake in Lagos, Nigeria: A multicentered randomized controlled trial. Vaccine, 38(42), 6600–6608. 10.1016/j.vaccine.2020.07.075

Manakongtreecheep, K. (2017). SMS-reminder for vaccination in Africa: Research from published, unpublished and grey literature. Pan African Medical Journal, 27(3), 23. 10.11604/pamj.supp.2017.27.3.12115

Mekonnen, Z. A., Gelaye, K. A., Were, M. C., Gashu, K. D., & Tilahun, B. (2021). Effect of mobile phone text message and call reminders on childhood vaccination timeliness and coverage: A systematic review and meta-analysis. JMIR mHealth and uHealth, 9(10), e27975. 10.2196/27975

Obi-Jeff, C., Garcia, C., Onuoha, O., Adewumi, F., David, W., Bamiduro, T., Aliyu, A. B., Labrique, A., & Wonodi, C. (2021). Designing an SMS reminder intervention to improve vaccination uptake in Northern Nigeria: A qualitative study. BMC Health Services Research, 21, 844. 10.1186/s12913-021-06728-2

Oladepo, O., Dipeolu, I. O., Olanrewaju, F. O., Osungbade, K. O., & Odebunmi, K. O. (2021). The effectiveness of mobile phone reminders on childhood immunization uptake in Oyo State, Nigeria. Journal of Public Health in Africa, 12(1), 1834. 10.4081/jphia.2021.1834

Yunusa, U., Idris, S. H., Adamu, H., & Baba, H. (2022). Effect of SMS reminder on immunization default among caregivers of under-five children in Zaria, Northwest Nigeria. Nigerian Journal of Paediatrics, 49(2), 82–88.

